# Standardized Data Elements for Patients with Acute Pulmonary Embolism: A Consensus Report from the Pulmonary Embolism Research Collaborative

**DOI:** 10.1101/2024.02.22.24303227

**Authors:** Kenneth Rosenfield, Terry R. Bowers, Christopher F. Barnett, George A. Davis, Jay Giri, James M. Horowitz, Menno V. Huisman, FESC, Beverley J. Hunt, Brent Keeling, Jeffrey A. Kline, Frederikus A. Klok, Stavros V. Konstantinides, Michelle T Lanno, Robert Lookstein, John M. Moriarty, Fionnuala Ní Áinle, Jamie L. Reed, Rachel P. Rosovsky, Sara M. Royce, Eric A. Secemsky, Andrew SP Sharp, Akhilesh K. Sista, Roy E. Smith, Phil Wells, Joanna Yang, Eleni M. Whatley

## Abstract

Recent advances in therapy and the promulgation of multidisciplinary pulmonary embolism teams (PERTs) show great promise to improve management and outcomes of acute pulmonary embolism (PE). However, the absence of randomized evidence and lack of consensus leads to tremendous variations in treatment and compromises the wide implementation of new innovations. Moreover, the changing landscape of healthcare, where quality, cost, and accountability are increasingly relevant, dictates that a broad spectrum of outcomes of care must be routinely monitored to fully capture the impact of modern PE treatment. We set out to standardize data collection in PE patients undergoing evaluation and treatment, and thus establish the foundation for an expanding evidence base that will address gaps in evidence and inform future care for acute PE. To do so, over 100 international PE thought leaders convened in Washington, DC in April 2022 to form the Pulmonary Embolism Research Collaborative (PERC™). Participants included physician experts, key members of the United States Food and Drug Administration (FDA), patient representatives, and industry leaders. Recognizing the multi-disciplinary nature of PE care, the Pulmonary Embolism Research Collaborative (PERC™) was created with representative experts from stakeholder medical subspecialties, including cardiology, pulmonology, vascular medicine, critical care, hematology, cardiac surgery, emergency medicine, hospital medicine, and pharmacology. A list of critical evidence gaps was composed with a matching comprehensive set of standardized data elements; these data points will provide a foundation for productive research, knowledge enhancement, and advancement of clinical care within the field of acute PE, and contribute to answering urgent unmet needs in PE management. Evidence produced through PERC™, as it is applied to data collection, promises to provide crucial knowledge that will ultimately produce a robust evidence base that will lead to standardization and harmonization of PE management and improved outcomes.

**CLINICAL PERSPECTIVE:** *What is new?:*

- Recent advances have increased options for treatment of acute pulmonary embolism, yet there remain wide variations in management due to the lack of a reliable evidence base upon which to base therapeutic decisions.
- The PERT Consortium^TM^ is a strong advocate of evidence based care for PE patients and therefore initiated the Pulmonary Embolism Research Collaborative (PERC^TM^) to establish a foundation for advancing high quality research and improving clinical care.
- A novel comprehensive set of standardized data elements is proposed for collection in patients with acute pulmonary embolism, to provide a foundation for expanding the evidence base and enhancing care.

*What are the clinical implications?:*

- Standardizing collection of data for acute pulmonary embolism will enable analyses that will inform optimal risk stratification, treatment, and follow-up of patients with pulmonary embolism, and provide evidence-based treatment algorithms that will improve outcomes.
- Registries created using the proposed standardized elements will enable benchmarking and quality assurance for clinicians caring for pulmonary embolism patients.
- Incorporation of comprehensive standardized data elements into FDA IDE trials will enable the Agency to better assess the safety and effectiveness of investigational devices.

## INTRODUCTION

Acute pulmonary embolism (PE) is a major source of cardiovascular morbidity and mortality and a leading cause of preventable in-hospital death in the US and around the world.^1–7^ Despite its high prevalence, clinical impact, and considerable public health implications, there remains a lack of consensus regarding what constitutes the optimal strategy for management of acute PE. The significant variability in the application of newer treatment modalities highlights the lack of consensus and need for standardization. Unlike myocardial infarction (MI) and stroke, for which robust evidence has led to consistent and standardized treatment, the absence of level I evidence regarding the management of PE has resulted in large variations in care. Further exacerbating this variability is the fact that PE is managed by several different specialties, each of which brings its own perspective and bias.

The most important reason for variability in treatment lies in the evidence gap that exists regarding optimal risk stratification and related risk-based management strategies, standardization of indication and application of interventional advanced PE treatment, and widely-accepted measures regarding outcomes of care for PE. Recent advances in treatment and availability of novel catheter directed techniques underscores the need for standardization of data collection in routine care, which is essential to understand their impact.

A core set of outcome measures for venous thromboembolism (VTE) was recently proposed by the International Consortium for Health Outcomes Measurement (ICHOM); however this set does not include relevant data elements regarding risk stratification and details of management decisions and is therefore insufficient to guide the much needed process of harmonization and standardization of PE management.^8, 9^ The Pulmonary Embolism Research Collaborative (PERC™) was conceived as a forum to develop a pragmatic core set of data elements, with common standards and definitions, that will serve as the foundation for data collection regarding the evaluation, treatment, outcomes, and follow up. The evidence generated by collecting such standardized data will enable analyses that will enhance clinical care, establish quality metrics, and promote research endeavors.

## METHODOLOGY

### PERC^TM^ composition

An international group of multidisciplinary experts in PE convened to work in collaboration with the United States Food and Drug Administration (FDA), patient representatives, healthcare organizations, and industry leaders, to explore gaps in recognition, diagnosis, treatment, and follow up of patients with acute PE. (**Figure 1**) PERC™ healthcare contributors represented the multidisciplinary character of PE care and included international representative experts from stakeholder medical subspecialties and scientific societies, including cardiology, pulmonology, vascular medicine, critical care, hematology, cardiac surgery, emergency medicine, hospital medicine, and pharmacology. Physician thought leaders were invited to participate based on their past contribution to PERT efforts as well as their involvement in contemporary trials for PE. Individuals invited to participate in PERC each obtained institutional approval to participate. FDA representatives received endorsement to participate on behalf of the Agency.

**Figure 1:**
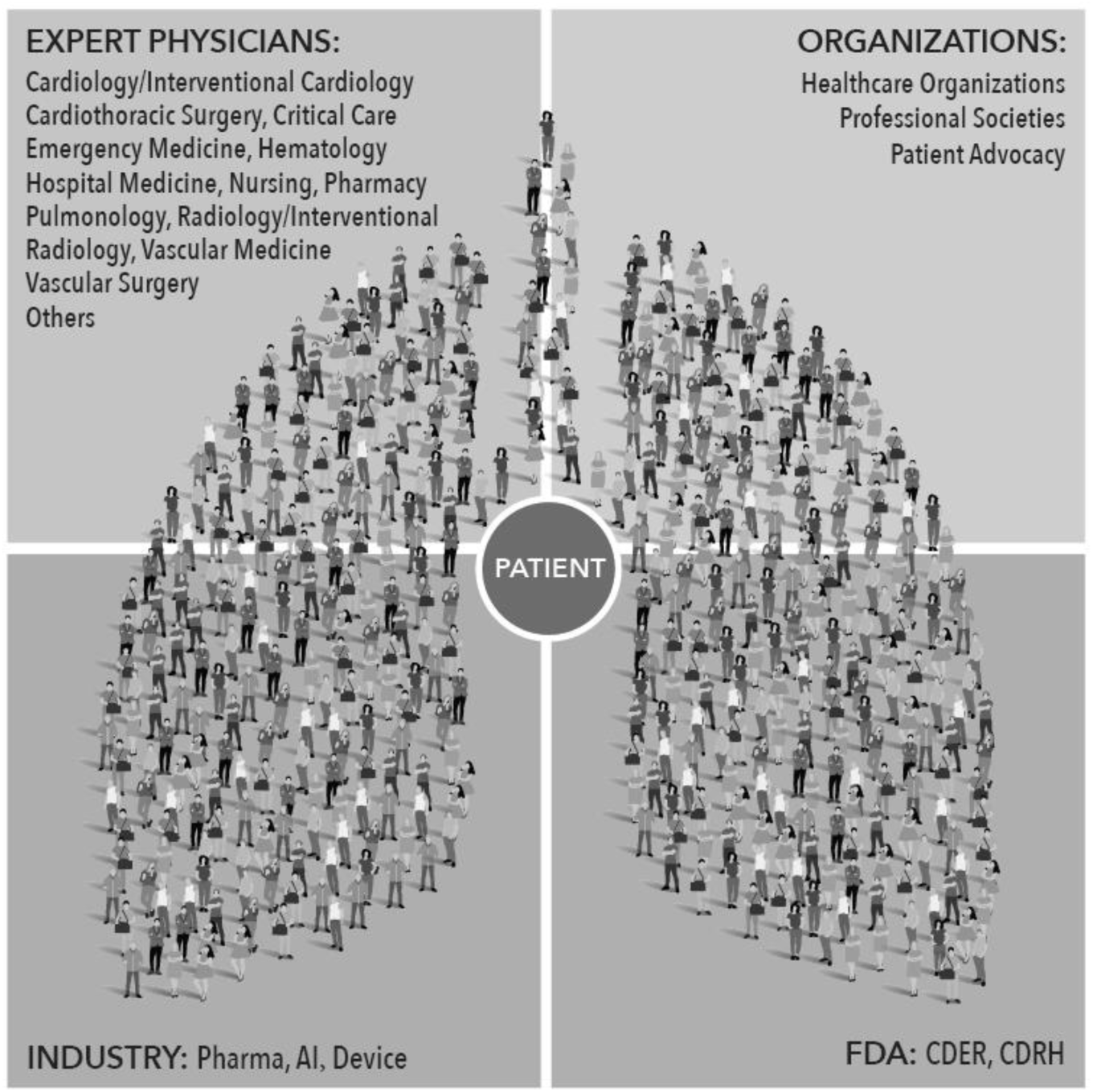
Multi-Disciplinary Experts contributing to Inaugural PERC™ Meeting

### PERC^TM^ objectives

The three objectives of PERC were: 1) to identify the current major evidence gaps that must be closed in order to better inform care providers and reduce variations in PE care; 2) to identify what data capture will be necessary to close those gaps; and 3) to develop consensus regarding a set of core set of data elements to be captured in routine care in all patients treated for PE. Ultimately, this initiative was to generate a comprehensive compendium of data elements pertaining to how patients are risk-stratified and treated, and to capture outcomes. Notably, PERC™ was not intended to establish recommended algorithms or guidelines for care. Ultimately, analyses derived from databases utilizing these standardized elements proposed by PERC™ will: a) establish the relative value and utility of current risk stratification tools; b) illuminate additional factors that may influence care and outcome, perhaps leading to creation of new tools for prognostication; c) enable more standardized and balanced assessment of outcomes using different therapeutic approaches; d) provide meaningful evidence to inform future guidelines and care algorithms.

### PERC^TM^ methodology and procedures

Five working groups (**Figure 2**), based on timing relative to patient presentation and intervention, were formulated before the meeting and leaders representing different disciplines were selected for each. Groups were charged with developing consensus around a set of data elements for capture pertaining to important knowledge gaps. In order to maximize productivity at the on-site PERC™ meeting, groups were asked to prepare their respective work product in advance. Accordingly, in the weeks leading up to the PERC™ meeting day, group leaders convened a series of workgroup virtual meetings. During the meeting, these proposed gaps in evidence and relevant data elements were discussed and finalized.

**Figure 2:**
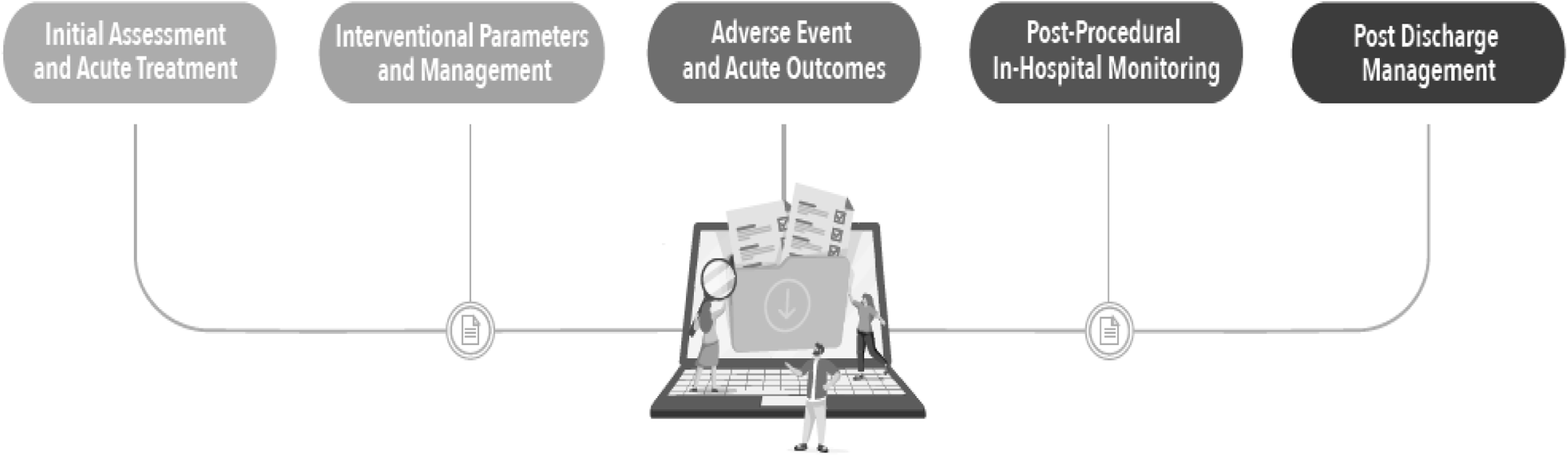
Data Element Collection throughout the 5 stages of the PE journey

### Establishment of list of data elements

Data elements throughout the entire “PE journey”, from presentation through discharge, and into follow up were considered in order to assess the various management strategies, decisions, and care rendered for acute PE, and the impact of these on outcomes. The timeline was divided into 5 distinct serial “events” representing milestones in the journey (**Figure 2**). Alternative (exploratory) elements not included in prior data collection efforts were considered, which could ultimately be relevant and predictive or influence outcomes, and therefore be important to capture. Elements were grouped into categories of “core” (i.e. absolutely necessary) and “enhanced” (i.e. beneficial to know for research and exploratory purposes). There was specific emphasis on identifying data elements that are pragmatic, such that they could be captured according to consensus definitions and at appropriate time points, with good ascertainment.

Practicable definitions that could be applied across institutions and easily understood by a broad professional audience were applied when available. Consideration was given to whether it was realistic to expect participating clinicians or institutions, during routine patient care, to collect the data element in a standardized manner. Evidence derived from the existing medical literature was used to justify final recommendations.

### Data sharing

An important concept in collection of data regarding PE from a broad base, as is expected in tools utilizing the PERC elements, is an agreement that data-sharing should be enabled to the extent that is legally and ethically possible. As such, users who share the common elements and definitions established by PERC will be encouraged to share de-identified data and thus enhance the overall dataset, enabling large-scale analyses that will increase the evidence base surrounding care of PE.

## RESULTS

Discussion in the working groups and during the PERC^TM^ meeting resulted in concentration on focused topics, including the current gaps in evidence and the need for specific data element capture.

### Gaps in Evidence

After discussion in the working groups and during the PERC^TM^ meeting, twelve crucial gaps in evidence were established as being the most important drivers of current variations in PE management and challenges that need to be overcome to improve the outcomes of care. (See **Table 1**). The gaps in evidence shown in were the driver of the selection and definition of data elements. An important unmet need addressed by the PERC™ experts was to clarify the roles of patient education, shared decision-making, and patient reported outcomes (PRO) within treatment paradigms.

**Table 1:**
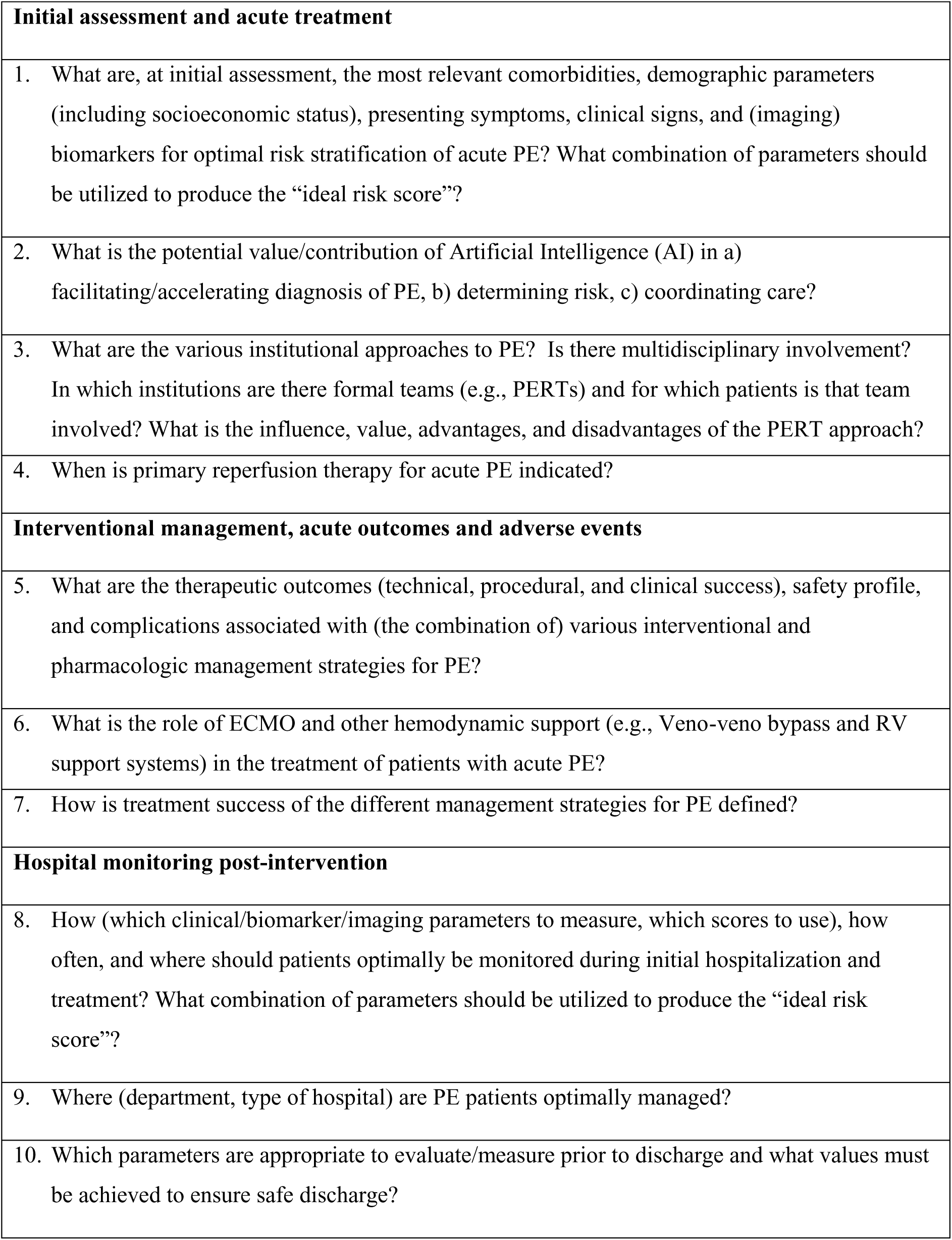
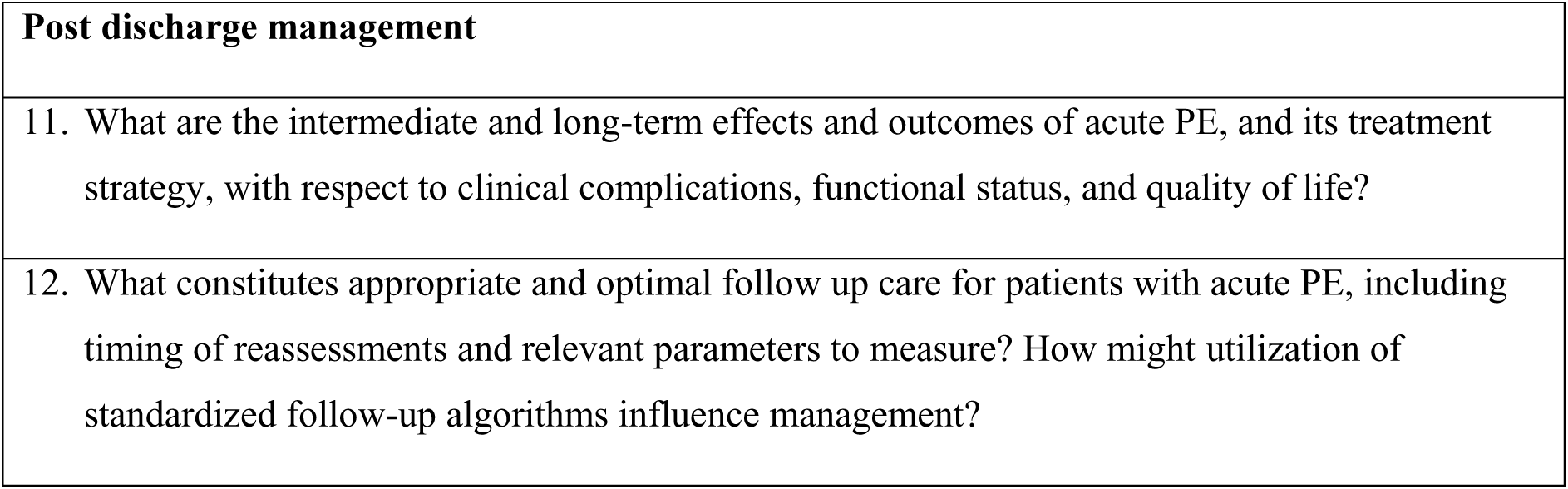
Overview of critical gaps in evidence per stage of the PE journey.

### List of Data Elements

During the PERC^TM^ meeting, 351 data elements were identified and defined. The final list of data elements listed fully in **Appendix 1** represent the ultimate output of the PERC™ meeting. For all data elements, the status (core versus enhanced) is indicated, as well as a definition, relevant side notes (e.g. relevant metadata standards) and/or a supporting reference where appropriate. The highlights of this list of data elements are detailed below.

The initial assessment includes common demographic parameters, risk factors and clinical details. Demographic elements determined to be of particular importance include sex assigned at birth (male, female, or other) and separate elements for race and ethnicity, based on recommendations from the Office of Minority Health.^10^ Standards for collection of race and ethnicity information are delineated on a prior document from the FDA entitled “Collection of Race and Ethnicity Data in Clinical Trials”^11^. Patient-level zip code is recommended as an important potential covariate for socioeconomic characterization, as is insurance status. This information is important to capture and evaluate, as health disparities in management of PE hospitalizations and PE follow-up, based on race^12, 13^, gender^14, 15^, and socioeconomic status^16^ have been reported.

Presenting clinical symptoms include those routinely endorsed by the patient on initial evaluation. Particular attention should be paid to the duration of PE symptoms (in days), as this may have significant interactions with the risk/benefit profiles of various PE therapeutics. Vital signs on admission and at various time points thereafter are relevant. Elements associated with hemodynamic compromise (e.g. cardiac arrest, hypotension requiring vasopressors, need for mechanical circulatory support) as well as respiratory compromise (e.g., requiring supplemental oxygen or mechanical ventilation) are captured. The recommended elements will enable calculations of various prognostic PE risk scores, including the sPESI score, Bova scores, National Early Warning Score and others.^17–20^ For that purpose, specific quantitative values of key laboratory tests are to be captured. For the same reason, the data elements also include details of initial imaging evaluations, including PE-protocol computed tomographic (CT) imaging and transthoracic echocardiography, performed either by a trained ultrasonographic technician/echocardiologist or as a point-of-care ultrasound (POCUS) by a physician or other qualified provider. Data elements associated with an escalation of care therapy, resulting in an (interventional) reperfusion procedure, should be captured, including all treatment pathways, along with specific parameters for each selected treatment approach. For all patients, typical procedural parameters, as well as baseline information that may inform the primary treatment decision, should be considered. These include, among others, date and time of PERT activation, the specialties involved in the team, and the presence of any known contraindication to anticoagulation and/or thrombolysis. For catheter-based therapies, such as catheter-directed thrombolysis (CDT) and catheter-based thrombectomy (CBT), complete device information, including either the unique device identifier (UDI) or brand, model and size, is pertinent. The exact location of treatment, access site information, and vascular closure information should be collected. For CDT, the drug type, dose, administration strategy (i.e., bolus vs infusion), and duration of treatment is essential. For CBT, whether the procedure entailed aspiration, mechanical disruption, or a combination is important. All elements related to the exact device type, treatment details, and drugs/dosages are considered core.

Procedure timing, specifically how to define the initiation of therapy and events related to a therapeutic intervention, is an important consideration and must be captured in order to enable accurate post-hoc analysis to evaluate and compare therapies. Given that specific data elements are being recommended to be conducted at various time points, the “start time” needs to be defined. This may be different for various therapies administered and, in the event that more than one therapy is administered, separate start times should be documented for each, rather than the whole “procedure.” For pharmaceutical treatments, including anticoagulation and systemic thrombolysis, the initiation of therapy is defined as the time when the drug is administered. For other device-based therapies, the initiation of therapy is defined as the time of venous access for the intervention. Specific focus is not placed on “door to catheter time,” though this should be calculable, as this has been reported that reduction of this time has a favorable impact on outcomes^21^. End of therapy is also captured, defined as follows:

- Oral anticoagulation– After administration of the first oral agent
- Parenteral anticoagulation– After injection of the first agent
- Systemic Thrombolysis – After the infusion is discontinued
- Catheter-based thrombolysis – After the completion of the lytic infusion
- Catheter-based thrombectomy – After leaving the angiographic suite
- Surgical thrombectomy – After leaving the operating room

Definitions for acute clinical success, which may differ based on the hemodynamic status of the patient, are summarized in **Table 2** and provide more detail to the definition of treatment success as suggested by the European Society of Cardiology.^22^ It is recognized that not achieving one or more of these definitions within 48 hours does not necessarily reflect that clinical improvement will not or has not partially been achieved (i.e., stabilized patient on ECMO support for >48 hours). However, to associate hemodynamic improvement with a therapeutic intervention requires proximity to the procedure (within 48 hours, by consensus of the group). It is recognized that not achieving one or more of these endpoints within 48 hours does not necessarily reflect that clinical improvement will not or has not partially been achieved (i.e., stabilized patient on ECMO support for >48 hours). However, to associate hemodynamic improvement with a therapeutic intervention requires proximity to the procedure (within 48 hours, by consensus of the group).

**Table 2:**
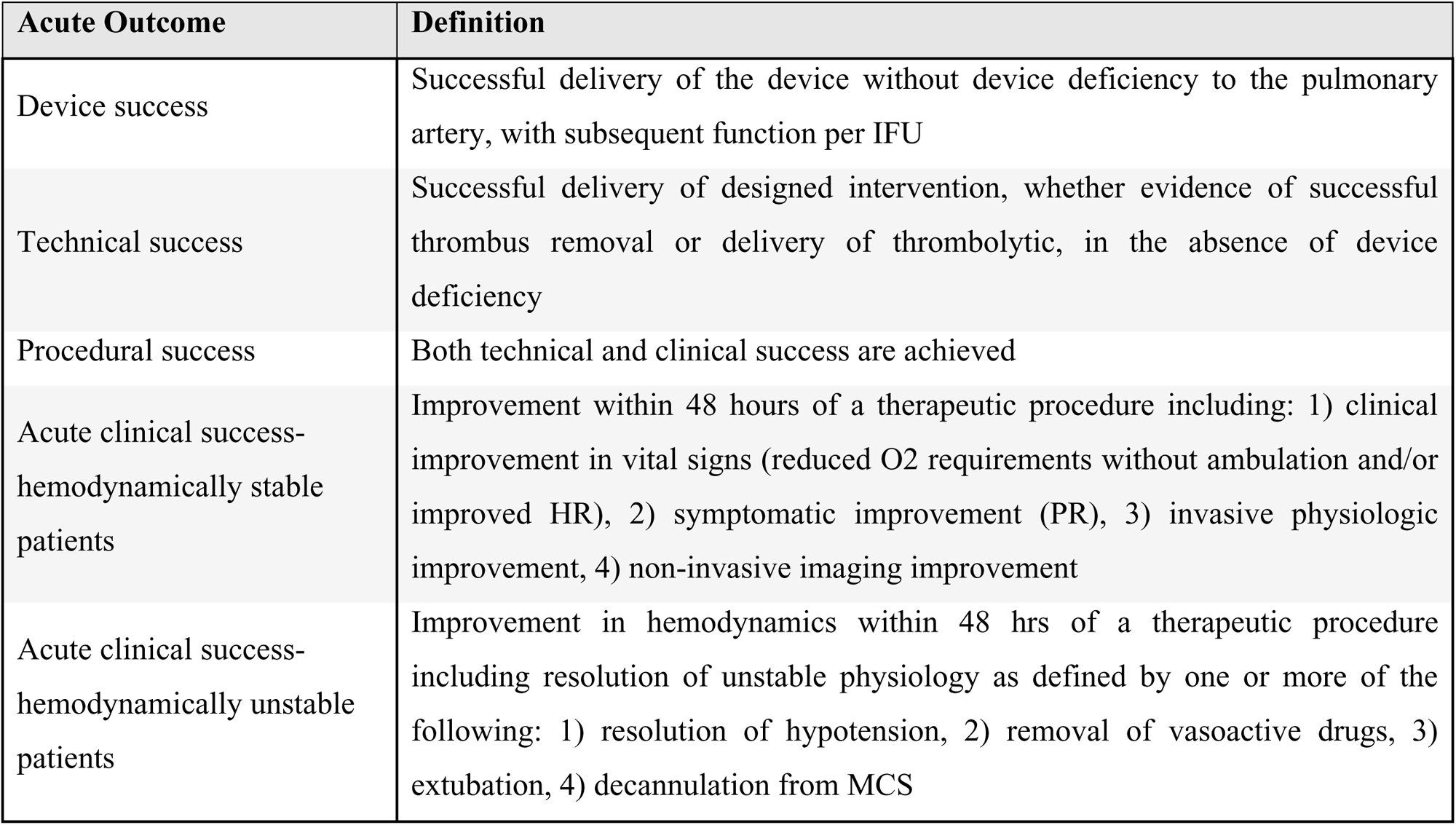
Definition of treatment success.

| Acute Outcome | Definition |
| --- | --- |
| Device success | Successful delivery of the device without device deficiency to the pulmonary artery, with subsequent function per IFU |
| Technical success | Successful delivery of designed intervention, whether evidence of successful thrombus removal or delivery of thrombolytic, in the absence of device deficiency |
| Procedural success | Both technical and clinical success are achieved |
| Acute clinical success-hemodynamically stable patients | Improvement within 48 hours of a therapeutic procedure including: 1) clinical improvement in vital signs (reduced O2 requirements without ambulation and/or improved HR), 2) symptomatic improvement (PR), 3) invasive physiologic improvement, 4) non-invasive imaging improvement |
| Acute clinical success-hemodynamically unstable patients | Improvement in hemodynamics within 48 hrs of a therapeutic procedure including resolution of unstable physiology as defined by one or more of the following: 1) resolution of hypotension, 2) removal of vasoactive drugs, 3) extubation, 4) decannulation from MCS |

Definitions for treatment failure and treatment-associated complications were agreed upon as well and illustrated in **Table 3**. Post-discharge management includes the collection of specific data elements post-discharge, such as adverse events (e.g. recurrent venous thromboembolism or bleeding), and hospital readmission rates at 30 days and 90 days after hospital discharge. Functional status and quality of life issues are considered part of this evaluation, even beyond the initial 3-month follow-up period, as well as the stepwise evaluation of patients for the development of chronic thromboembolic pulmonary hypertension (CTEPH) or chronic thromboembolic pulmonary disease (CTEPD).^23–27^

**Table 3:** Definition of treatment failure and complications of treatment.

| Adverse Event | Definition |
| --- | --- |
| Major bleeding | ISTH definition |
| Hemoptysis | If a patient coughs up blood with sputum or has frank bleeding into the airway. If present, classify into moderate or major |
| Moderate hemoptysis | Less than 50 mL of blood without the need for additional intervention |
| Major hemoptysis | Greater than 50 mL of blood with or without additional intervention |
| Impending respiratory arrest | A patient with intact neurologic function becomes agitated, confused and struggles to breathe. Usually associated with tachycardia, diaphoresis and intercostal or sternoclavicular retractions |
| Device malfunction | Failure of a medical device to perform in accordance with its intended purpose when used in accordance with the instructions for use |
| Device user error | User action or lack of user action while using the medical device that leads to a different result than that intended by the manufacturer or expected by the user |
| Device Inadequacy of IFU | Insufficient or misleading information in the instructions for use or labeling |
| Device fracture/component separation | Undesirable damage to a medical device involving the loss of structural integrity, such as a partial or full thickness crack in the device materials, or loss of adherence between device materials, resulting in an undesirable separation, breaking apart, or detachment of some part or piece(s) of the device |
| Access site complication | Any persistent bleeding or hematoma in conjunction with the need for transfusion, or access-site vascular injury requiring intervention (e.g. surgical or endovascular repair) |
| Clinical deterioration | Defined as one in the following: 1) need for cardiopulmonary resuscitation, 2) systolic blood pressure < 90 mm Hg for at least 15 min, 3) drop in systolic blood pressure by at least 40 mm Hg for at least 15 minutes with signs of end-organ |
|  | hypoperfusion (cold extremities or low urinary output < 30 mL/h or mental confusion), 4) need for catecholamine administration to maintain adequate organ perfusion and a systolic blood pressure of > 90 mm Hg (including dopamine at the rate of > 5 microgram/kg per minute), 5) unplanned endotracheal intubation, 6) unexpected requirement for mechanical ventilation, 7) emergency surgical embolectomy |
| Device-related death at 48 hours | Death directly related to the device, including from the following: 1) vascular or cardiovascular injury, 2) device malfunction, 3) device-induced cardiac arrhythmia, 4) worsening pulmonary or right heart function (exclusive of worsening from recurrent PE) |

Lastly, the data elements include indicators of information having been provided for education of the patient and his/her family, in particular regarding the use of anticoagulant drugs.

## DISCUSSION

Large gaps in the evidence base regarding management of PE have resulted in substantial variations in care. Identification and capture of appropriate data elements during the journey of the PE patient, from presentation to discharge and into follow-up, will be critical to expansion of the evidence base and represent the first step towards a more pragmatic and evidence-based approach to patient care in this rapidly evolving field. The current document establishes a baseline of relevant data elements, generated by consensus of the Pulmonary Embolism Research Collaborative (PERC^TM^) in conjunction with FDA, that are appropriate to collect during the PE patient journey. Elements are included on the basis of their known or potential effect on outcome and are designated as either “core” or “enhanced”. The core set represents the minimum necessary elements for capture, while the enhanced set includes additional parameters that may be useful to inform future understanding of PE and influence treatment strategies. Analysis of these data will enlighten the medical community and provide guidance regarding current best practices, and will serve as the foundation for improving therapies moving forward. While establishing a standardized set of data elements was the main objective of the PERC™, another key objective of the initiative was for the expert collaborators to collectively identify the shortcomings and data gaps in PE care.

During the course of the PERC™ deliberations, in addition to identifying specific elements worthy of capture, several important considerations were identified that affect PE data collection. Addressing each of these will be important as part of the effort to expand the PE evidence base and close the knowledge gap regarding optimal management.

### Threshold for Activation and Data Capture

The “spectrum of PE”, or the varying degrees in which this disease presents, is an acknowledged phenomenon and is a function of many variables. The threshold for various treatments, based on this spectrum, is not well defined. Similarly, the threshold for full PERT activation is not standardized in regard to this spectrum. Indeed, the significant variations in care were the stimulus for promoting the team-based concept of PE care (PERTs) and are one of the most compelling reasons for collecting and analyzing patterns of PE care in a registry. Acknowledging the variations in presentation and severity of PE, the data elements recommended for capture herein may not be necessary or appropriate for every patient with acute PE.

### Statistical Analysis

Data acquired through registries and other sources using the PERC template and definitions will be subjected to rigorous review as a part of all subsequent analyses. Missingness, which plagues many registry databases, will be addressed as part of this review. The pragmatism of fully capturing data elements will be affected by many factors, including then number and complexity of the elements identified, the resources available at the point of collection, and the level of ascertainment required by the respective registry or collection tool. The data elements recommended for capture within this document represent a comprehensive list of those that were identified as minimally necessary elements (i.e. core) or those which provide for more detailed analysis or are exploratory regarding their impact on outcome (i.e. enhanced). Both core and enhanced data may be useful to inform future understanding of PE and influence treatment strategies. It is acknowledged that these constitute a long list of elements, which may be challenging to capture fully and comprehensively. It is also understood that, as the list of elements for capture increases, the potential for increase in missing data also exists. Appropriate data ascertainment is important since significant missing data will hinder the data evaluation and analytic process. Thus, the intention of this document is to emphasize capture of the core elements. Data quality and integrity and the value of the subsequent statistical analyses will rely upon identification of the most critical (i.e. essential) elements and sufficient ascertainment thereof.

### Data Collection Timepoints

The timing and, more importantly, the frequency with which data elements should be captured for input into any registry or clinical trial database is an important consideration. PE patients represent a wide spectrum of severity and, therefore, the level of monitoring and data capture will vary as well. For example, a patient with a recent cardiac arrest would be receiving real-time electronic monitoring with continuous recording of data from oxygen saturation probes, arterial blood pressure recording lines, and respiratory rates. Whereas, a patient who is stable and minimally symptomatic may be admitted to a standard medical floor and only have clinical observations documented every six hours. It would not be appropriate to expect those respective frequencies to be required for capture and, on the other hand, a less rigorous approach might fail to capture important and relevant information. To strike a balance, it was felt that observations on all patients should be recorded and inputted to registries at a minimum of once per day, with more intense requirements for those at the more critical end of the spectrum. Of course, this is conjecture and the ideal timepoints are yet to be understood. An ability to interface national PE registries with electronic medical records (EMR) may be helpful in this respect, as software bridges could be written that allow automated extraction of clinical observations data from the EMR source. This remains to be further explored, as ongoing expansion of the evidence base sheds light on optimal timepoints and intervals for data collection.

### Use of AI for Data Capture

Artificial Intelligence (AI) is playing an increasingly important role in facilitating the diagnosis and management of acute PE. Machine learning algorithms can expedite and enhance the detection of PE on CT scans, as well as the presence of echocardiographic findings such as RV strain. Further, AI programs can integrate clinical data to define risk scores and facilitate decision-making. Ultimately, AI may not only inform the clinical team and guide management but may also suggest new management paradigms based on input from outcomes combined with computer mediated “deep learning.”

Current AI programs access raw data from imaging modalities and subject those data to analysis to define presence of a PE and its characteristics. AI programs are now establishing communication pathways with electronic medical records, allowing for assimilation of patient data (including laboratory values, clinical information, and test results). Integration of these data, when subjected to AI tools such as machine and deep learning, will provide a more comprehensive analysis of patient risk, clinical status, and optimal treatment.

AI will be a powerful tool to optimize data capture and provide a more robust evidence base for PE. During the process of accessing and organizing raw data from imaging modalities and the electronic medical record, these programs can be utilized to automatically populate PE databases. By enabling data collection “in the background,” AI will provide more standardization of data collection, reduce subjectivity, ensure more consistency in interpretation, and reduce “missingness” (enhance ascertainment). Beyond this, deep (“cognitive”) learning extrapolated from these data will add a new dimension to the evidence base and ultimately enhance the treatment paradigms for PE, through more accurate calculation of existing risk scores, creation of novel, more comprehensive risk calculators, and establishment of increasingly effective care pathways.

### Partnership with FDA

FDA has been an active participant in this project and shares common goals of optimizing care, improving outcomes, and ensuring safety and efficacy in the care of PE patients. The challenges facing treating clinicians are similar to the questions that regulatory experts grapple with when evaluating the risk-benefit profile of new PE devices and treatment strategies. FDA is committed to least burdensome approaches^28^ to marketing applications. Better understanding of decision-making surrounding escalation and use of advance therapies, as well as the relative effectiveness of various treatment strategies in different PE populations, can inform regulatory strategies, potentially streamline the availability of effective new treatments, and foster innovation in the care of the PE patient. Ongoing participation of regulatory agencies in PERC™, coupled with an associated robust registry, provides an opportunity to assist in regulatory decision making. This supports the FDA’s priority to advance the use of real-world evidence (RWE)^29, 30^.

## CONCLUSION

The PERC™ initiative was designed to identify gaps in the knowledge base regarding care of pulmonary embolism and to establish a standardized set of data elements to collect in patients with PE. Utilization of the elements identified and defined by PERC™ within registries and clinical trials, with appropriate data-sharing arrangements, will enable coalescence of data to perform large-scale analyses that will expand the evidence base for PE and ultimately inform best practices. Engagement of multispecialty physicians, device manufacturers, pharmacological experts, patient advocates, as well as regulatory representatives from both device and drug agencies in the PERC™ process enabled a broad-based understanding of the evidence gaps. PERC™ has set the stage to address these evidence gaps and promote progress in PE care.

## Data Availability

All data are original

## Disclaimer

This article reflects the consensus views of the writing group and does not necessarily represent the practices, policies, requirements or recommendations of the FDA. Further, the use of the word “required”, “must” or “should” in the document is not intended to indicate an FDA requirement.

## Disclosures

Dr. Rosenfield reports consult fees from Abbott Vascular, Angiodynamics, Inc., Auxetics, Becton-Dickinson, Boston Scientific, Inc., Contego, Imperative Care, Johnson & Johnson, Biosense Webster, Medtronic, Neptune Medical, Philips, Surmodics, and Terumo, Board Member for The PERT Consortium™, and stock or stock options in Access Vascular, Aerami, Althea Medical, Auxetics, Contego, Endospan, Imperative Care, Innova Vascular, InspireMD, JanaCare, Magneto, MedAlliance, Neptune Medical, Orchestra, Prosomnus, Sealonix, Shockwave, Skydance, Summa Therapeutics, Thrombolex, Valcare, Vantis Vascular, Vasorum, and Vumedi.

Dr. Bowers reports consulting fees from Truvic, Inc., lecture fees from Janssen and BMS-Pfizer, payment for expert testimony, participation on Advisory Board of Truvic, Inc.

Dr. Barnett reports payment for expert testimony.

Dr. Davis reports travel support for attending the Pulmonary Embolism Research Collaborative meeting.

Dr. Giri reports research support from Boston Scientific, Inc. and Inari Medical, consulting fees from Boston Scientific, Inc., Cordis, and Abbott Vascular, support for attending meetings from Inari Medical, participation on an Advisory Board for Angiodynamics, Inc. and Boston Scientific, Inc., participation in leadership at SCAI and The PERT Consortium™, and stock or stock options in Endovascular Engineering.

Dr. Horowitz reports research support from Inari Medical, consultation and personal lecture fees from Inari Medical and Penumbra, Inc., and support for attending meetings from Inari Medical.

Professor Dr. Huisman reports no conflicts of interest.

Professor Dr. Hunt reports no conflicts of interest.

Dr. Keeling reports consulting fees from Angiodynamics, Inc., Penumbra, Inc., Viz.ai, and Dexcom.

Dr. Kline reports no conflicts of interest.

Professor Dr. Klok reports institutional research support from Bayer, Bristol-Myers Squibb, Actelion, Boston Scientific, Inc, Leo Pharma, The Netherlands Organisation for Health Research and Development, The Dutch Thrombosis Association, The Dutch Heart Foundation, and the Horizon Europe program, service on the Board of the Rembrandt Institute of Cardiovascular Science, Chair of the Board of the Dutch Society of Vascular Medicine, nucleus member of the ESC work group on pulmonary circulation and RV function, nucleus member of ISTH SSC diagnostic and predictive variables, Chair of the Board of the Dutch Thrombosis Network, and medical lead Anticoagulation Clinic Leiden.

Professor Dr. Konstantinides reports personal and institutional consulting fees from Bayer AG, Daiichi-Sankyo, and Boston Scientific, Inc., and personal lecture fees from Bayer AG, Daiichi-Sankyo, Pfizer – Bristol-Myers Squibb, and Boston Scientific, Inc.

Ms. Lanno reports no conflicts of interest.

Dr. Lookstein reports consulting fees from Penumbra, Inc., Boston Scientific, Inc., Medtronic, Cordis, Becton Dickinson, Abbott Vascular, Neptune Medical, Imperative Care, and Aidoc, payment for expert testimony, participation on Advisory Board for Magneto, Boston Scientific, Inc., Medtronic, and Trireme, leadership roles at The PERT Consortium™, CLI Global Society, and Society of Interventional Radiology, stock or stock options in Imperative Vascular, Thrombolex, Innova Vascular, and Summa Vascular.

Dr. Moriarty reports research support from Angiodynamics, Inc. and Penumbra, Inc., consulting fees from Angiodynamics, Inc, Penumbra Inc., Inquis Medical, Innova Vascular, Boston Scientific Inc., Auxetics, and Retriever Medical, and participation on the data safety monitoring board for Penumbra, Inc.

Professor Dr. Ní Áinle reports institutional research support from Sanofi, Daiichi-Sankyo, the Irish Health Research Board, Bayer, and Boston Scientific, Inc., patents pending for European Application No. 20166826.6, past member of World Thrombosis Day International Steering Committee, Director at Large and Member, INVENT Council, Co-Director, InVite, Co-Founder and member of Executive Committee of VTE Ireland, Advisory Committee for ACHM, Member of “Preventing venous thromboembolism in hospitals” Collaborative Advisory Group, Board Member, Thrombosis Ireland Patient Organization and Charity, and National Lead, Irish VTE Patient Safety Programme.

Ms. Reed reports no conflicts of interest.

Dr. Rosovsky reports research support from BMS and Janssen, consulting fees from Abbott, BMS, Dova, Inari Medical, Janssen, and Penumbra, Inc., participation on the data safety monitoring board for the Pepper Trial and Harvard Cancer Center, unpaid President-Elect of the PERT Consortium and unpaid member of ASH Committee on Quality.

Dr. Royce reports no conflicts of interest.

Dr. Secemsky reports research support from NIH/NHLBINIH/NHBLI K23HL150290, Food & Drug Administration, BD, Boston Scientific, Inc., Cook Medical, CSI, Laminate Medical, Medtronic, and Philip, consulting fees and personal lecture fees from Abbott, Bayer, BD, Boston Scientific, Inc., Cook, CSI, Inari Medical, Medtronic, Philips, Shockwave, and VentureMed, ad participation on data safety monitoring boards for LIFE BTK, BIOMIMCS3D, HI-PEITHO, and Transcend.

Professor Dr. Sharp reports consulting and lecture fees from Medtronic, Boston Scientific, Inc., Philips, Recor Medical, and Penumbra, Inc.

Dr. Sista reports research support from NIH/NHLBI as principal investigator of U34 and UG3 awards, unpaid participation on data safety monitoring boards for Angiodynamics, Inc. and Thrombolex, Inc., and participation on the Board of Directors for the Society of Interventional Radiology.

Dr. Smith reports no conflicts of interest.

Dr. Wells reports participation on the data safety monitoring board for Anthos. Ms. Yang reports no conflicts of interest.

Dr. Whatley reports no conflicts of interest.

## ABBREVIATIONS

AI: Artificial Intelligence
AC: Anticoagulation; Anticoagulant
CBT: Catheter-based Thrombectomy
CDL: Catheter-directed Thrombolysis
CDER: Center for Drug Evaluation and Research
CDRH: Center for Devices and Radiological Health
CPET: Cardiopulmonary Exercise Testing
CTEPD: Chronic Thromboembolic Pulmonary Disease (also referred to as CTED)
CTEPH: Chronic Thromboembolic Pulmonary Hypertension
ECMO: Extracorporeal Membrane Oxygenation
EMR: Electronic Medical Record
ESC: European Society of Cardiology
FDA: Food and Drug Administration
MCS: Mechanical Circulatory Support
PE: Pulmonary Embolism
PERC: Pulmonary Embolism Research Collaborative
PERT: Pulmonary Embolism Response Team
PRO: Patient Reported Outcome
VTE: Venous Thromboembolism

## ACKNOWLEDGEMENTS

We would like to acknowledge the contributions from all attendees at the inaugural PERC™™ meeting held in Washington DC on April 22, 2022 (**Appendix 2**).

## APPENDICES

## Appendix 1 Core and Enhanced Data Elements for Collection in Acute PE Patients

*See attached Excel spreadsheet*

## Appendix 2 Inaugural PERC^TM^ Meeting Contributors

*See attached Word document*

## REFERENCES

1. Sedhom R, Megaly M, Elbadawi A, Elgendy IY, Witzke CF, Kalra S, et al. Contemporary national trends and outcomes of pulmonary embolism in the united states. Am J Cardiol. 2022;176:132–138

2. Benjamin EJ, Virani SS, Callaway CW, Chamberlain AM, Chang AR, Cheng S, et al. Heart disease and stroke statistics-2018 update: A report from the american heart association. Circulation. 2018;137:e67–e492

3. Heit JA, Spencer FA, White RH. The epidemiology of venous thromboembolism. J Thromb Thrombolysis. 2016;41:3–14

4. Smith SB, Geske JB, Kathuria P, Cuttica M, Schimmel DR, Courtney DM, et al. Analysis of national trends in admissions for pulmonary embolism. Chest. 2016;150:35–45

5. Barco S, Mahmoudpour SH, Valerio L, Klok FA, Munzel T, Middeldorp S, et al. Trends in mortality related to pulmonary embolism in the european region, 2000-15: Analysis of vital registration data from the who mortality database. Lancet Respir Med. 2020;8:277–287

6. Barco S, Valerio L, Ageno W, Cohen AT, Goldhaber SZ, Hunt BJ, et al. Age-sex specific pulmonary embolism-related mortality in the USA and canada, 2000-18: An analysis of the who mortality database and of the cdc multiple cause of death database. Lancet Respir Med. 2021;9:33–42

7. Huisman MV, Barco S, Cannegieter SC, Le Gal G, Konstantinides SV, Reitsma PH, et al. Pulmonary embolism. Nat Rev Dis Primers. 2018;4:18028

8. Gwozdz AM, de Jong CMM, Fialho LS, Likitabhorn T, Sossi F, Jaber PB, et al. Development of an international standard set of outcome measures for patients with venous thromboembolism: An international consortium for health outcomes measurement consensus recommendation. Lancet Haematol. 2022;9:e698–e706

9. de Jong CMM, Rosovsky RP, Klok FA. Outcomes of venous thromboembolism care: Future directions. J Thromb Haemost. 2023;21:1082–1089

10. U.S. Department of Health and Human Services Office of Minority Health. Homepage. 2022

11. U.S. Food and Drug Administration Communication. Collection of race and ethnicity data in clinical trials. 2016

12. Martin KA, McCabe ME, Feinglass J, Khan SS. Racial disparities exist across age groups in illinois for pulmonary embolism hospitalizations. Arterioscler Thromb Vasc Biol. 2020;40:2338–2340

13. Phillips AR, Reitz KM, Myers S, Thoma F, Andraska EA, Jano A, et al. Association between black race, clinical severity, and management of acute pulmonary embolism: A retrospective cohort study. J Am Heart Assoc. 2021;10:e021818

14. Pribish AM, Beyer SE, Krawisz AK, Weinberg I, Carroll BJ, Secemsky EA. Sex differences in presentation, management, and outcomes among patients hospitalized with acute pulmonary embolism. Vasc Med. 2020;25:541–548

15. Agarwal S, Clark D, 3rd, Sud K, Jaber WA, Cho L, Menon V. Gender disparities in outcomes and resource utilization for acute pulmonary embolism hospitalizations in the united states. Am J Cardiol. 2015;116:1270–1276

16. Wadhera RK, Secemsky EA, Wang Y, Yeh RW, Goldhaber SZ. Association of socioeconomic disadvantage with mortality and readmissions among older adults hospitalized for pulmonary embolism in the united states. J Am Heart Assoc. 2021;10:e021117

17. Aujesky D, Obrosky DS, Stone RA, Auble TE, Perrier A, Cornuz J, et al. Derivation and validation of a prognostic model for pulmonary embolism. Am J Respir Crit Care Med. 2005;172:1041–1046

18. Bova C, Sanchez O, Prandoni P, Lankeit M, Konstantinides S, Vanni S, et al. Identification of intermediate-risk patients with acute symptomatic pulmonary embolism. Eur Respir J. 2014;44:694–703

19. RCoPo L. National early warning score (news): Standardising the assessment of acute-illness severity in the nhs — report of a working party. Royal College of Physicians. 2012

20. Bavalia R, Stals MAM, Mulder FI, Bistervels IM, Coppens M, Faber LM, et al. Use of the national early warning score for predicting deterioration of patients with acute pulmonary embolism: A post-hoc analysis of the years study. Emerg Med J. 2023;40:61–66

21. Rawal A, Ardeshna D, Hesterberg K, Cave B, Ibebuogu UN, Khouzam RN. Is there an optimal “door to cath time” in the treatment of acute pulmonary embolism with catheter-directed thrombolysis? Ann Transl Med. 2019;7:419

22. Pruszczyk P, Klok FA, Kucher N, Roik M, Meneveau N, Sharp ASP, et al. Percutaneous treatment options for acute pulmonary embolism: A clinical consensus statement by the esc working group on pulmonary circulation and right ventricular function and the european association of percutaneous cardiovascular interventions. EuroIntervention. 2022;18:e623–e638

23. Boon G, Barco S, Bertoletti L, Ghanima W, Huisman MV, Kahn SR, et al. Measuring functional limitations after venous thromboembolism: Optimization of the post-vte functional status (pvfs) scale. Thrombosis research. 2020;190:45–51

24. Klok FA, Cohn DM, Middeldorp S, Scharloo M, Buller HR, van Kralingen KW, et al. Quality of life after pulmonary embolism: Validation of the pemb-qol questionnaire. J Thromb Haemost. 2010;8:523–532

25. Delcroix M, Torbicki A, Gopalan D, Sitbon O, Klok FA, Lang I, et al. Ers statement on chronic thromboembolic pulmonary hypertension. Eur Respir J. 2021;57

26. Humbert M, Kovacs G, Hoeper MM, Badagliacca R, Berger RMF, Brida M, et al. 2022 esc/ers guidelines for the diagnosis and treatment of pulmonary hypertension. Eur Heart J. 2022;43:3618–3731

27. Morris TA, Fernandes TM, Channick RN. Evaluation of dyspnea and exercise intolerance after acute pulmonary embolism. Chest. 2023;163:933–941

28. U.S. Food and Drug Administration. The least burdensome provisions: Concept and principles. 2019

29. U.S. Food and Drug Administration. Use of real-world evidence to support regulatory decision-making for medical devices. 2017

30. U.S. Food and Drug Administration. Real-world data: Assessing registries to support regulatory decision-making for drug and biological products - guidance for industry. 2021

31. Stein PD, Matta F, Goldman J. Obesity and pulmonary embolism: The mounting evidence of risk and the mortality paradox. Thrombosis research. 2011;128:518–523

32. Gauthier K, Kovacs MJ, Wells PS, Le Gal G, Rodger M, investigators R. Family history of venous thromboembolism (vte) as a predictor for recurrent vte in unprovoked vte patients. J Thromb Haemost. 2013;11:200–203

33. Konstantinides SV, Meyer G, Becattini C, Bueno H, Geersing GJ, Harjola VP, et al. 2019 esc guidelines for the diagnosis and management of acute pulmonary embolism developed in collaboration with the european respiratory society (ers): The task force for the diagnosis and management of acute pulmonary embolism of the european society of cardiology (esc). Eur Respir J. 2019;54

34. Budd AC, Rhodes M, Forster AJ, Noghani P, Carrier M, Wells PS. Prescribing patterns and outcomes of venous thromboembolism prophylaxis in hospitalized medical and cancer patients: Observations from the ottawa hospital. Thrombosis research. 2021;197:144–152

35. Garcia D, Akl EA, Carr R, Kearon C. Antiphospholipid antibodies and the risk of recurrence after a first episode of venous thromboembolism: A systematic review. Blood. 2013;122:817–824

36. Rodger MA, Kahn SR, Wells PS, Anderson DA, Chagnon I, Le Gal G, et al. Identifying unprovoked thromboembolism patients at low risk for recurrence who can discontinue anticoagulant therapy. CMAJ. 2008;179:417–426

37. de Winter MA, van Es N, Buller HR, Visseren FLJ, Nijkeuter M. Prediction models for recurrence and bleeding in patients with venous thromboembolism: A systematic review and critical appraisal. Thrombosis research. 2021;199:85–96

38. Prins MH, Lensing AWA, Prandoni P, Wells PS, Verhamme P, Beyer-Westendorf J, et al. Risk of recurrent venous thromboembolism according to baseline risk factor profiles. Blood Adv. 2018;2:788–796

39. Bates SM, Ginsberg JS. Clinical practice. Treatment of deep-vein thrombosis. N Engl J Med. 2004;351:268–277

40. Rosano GMC, Rodriguez-Martinez MA, Spoletini I, Regidor PA. Obesity and contraceptive use: Impact on cardiovascular risk. ESC Heart Fail. 2022

41. Schulman S, Kearon C, Subcommittee on Control of Anticoagulation of the S, Standardization Committee of the International Society on T, Haemostasis. Definition of major bleeding in clinical investigations of antihemostatic medicinal products in non-surgical patients. J Thromb Haemost. 2005;3:692–694

42. Quigley N, Gagnon S, Fortin M. Aetiology, diagnosis and treatment of moderate-to-severe haemoptysis in a north american academic centre. ERJ Open Res. 2020;6

43. Meyer G, Vicaut E, Danays T, Agnelli G, Becattini C, Beyer-Westendorf J, et al. Fibrinolysis for patients with intermediate-risk pulmonary embolism. N Engl J Med. 2014;370:1402–1411

44. The Joint Commission. National patient safety goal for anticoagulant therapy. 2018:1–4

45. Konstantinides SV, Meyer G, Becattini C, Bueno H, Geersing GJ, Harjola VP, et al. 2019 esc guidelines for the diagnosis and management of acute pulmonary embolism developed in collaboration with the european respiratory society (ers). Eur Heart J. 2020;41:543–603

46. Gaddh M, Rosovsky RP. Venous thromboembolism: Genetics and thrombophilias. Semin Respir Crit Care Med. 2021;42:271–283

47. Khair RM, Nwaneri C, Damico RL, Kolb T, Hassoun PM, Mathai SC. The minimal important difference in borg dyspnea score in pulmonary arterial hypertension. Ann Am Thorac Soc. 2016;13:842–849

